# Implementation of a Rapid RT-LAMP Saliva-based SARS-CoV-2 Testing Program in the Workplace

**DOI:** 10.1101/2022.01.15.22269318

**Authors:** Bradley W.M. Cook, Kaitlyn Kobasa, Marielou Tamayo, Natasha Theriault, Diane Gordon Pappas, Steven S. Theriault

**Author notes:** These authors have contributed equally to this work (Kaitlyn Kobasa, Marielou Tamayo, Natasha Theriault).

## Abstract

Rising SARS-CoV-2 cases, testing delays and the risk of pre-symptomatic and asymptomatic transmission provided the impetus for an in-house rapid testing program. Employees and their household contacts were encouraged to self-collect saliva samples which were pooled for routine testing using an established colorimetric reverse transcription loop-mediated isothermal amplification (RT-LAMP) assay. In brief, individual or a maximum of four saliva samples were pooled, heat-inactivated to render microorganisms, especially SARS-CoV-2, non-infectious prior to being added to RT-LAMP assay tubes containing either human sample control gene, RNase P or a region of the SARS-CoV-2 gene, ORF1ab. During the second wave of SARS-CoV-2 infections in November 2020, two samples from an employee and a member of their household tested positive via RT-LAMP within two days of each other. A delayed clinical qRT-PCR test confirmation of both individuals 5 days later underscores the power of routine rapid testing with with-in-the-hour turnaround times. Workplace rapid testing programs using RT-LAMP are flexible in their design, have a reduced cost compared to qRT-PCR, may involve non-invasive self-saliva collection for increased safety for the testing personnel, and can be performed with minimal training.

## Introduction

Quantitative reverse-transcription-polymerase chain reaction (qRT-PCR) is recognized as the gold standard diagnostic test to determine if an individual is infected with SARS-CoV-2. Samples are obtained from a nasopharyngeal swab or the less invasive, nasal swab. This test strategy requires costly and sometimes supply limited consumables, hours of skilled labor, and expensive machinery for testing and analysis [1]. All of which are typically not available outside of clinical settings or an established academic laboratory. During the summer of 2020 in Manitoba, Canada the demand for qRT-PCR testing led to processing delays and extended turnaround timesup to several days for results [2]–[4]. Long turnaround times would exacerbate community spread as testing is voluntary and positive results would be skewed towards symptomatic individuals [5], in Manitoba more than two-thirds submitted for testing were symptomatic (summarized in Figure 1). This bias meant that 20-59% of infected individuals which are either pre-symptomatic or asymptomatic were unobserved by public health authorities and may have led to further transmission [6], [7]. The public health impact of such delays, necessitate the use of cheap, easy-to-manufacture, scalable and affordable rapid testing options which can aid in the ability to pre-screen for potentially contagious individuals. Rapid tests can be performed without highly trained personnel or complicated machinery (end point is detected visually) and are amenable for rapid daily-use to detect contagious individuals [1]. Reverse transcription loop-mediated isothermal amplification (RT-LAMP) is a rapid testing technique which amplifies target gene(s) resulting in continual amplification of stem-loop DNA structures at a single temperature [8]. The assay for SARS-CoV-2 detection used in this screening program was previous optimized by Yang et. al. 2020, for use with acidic saliva samples and a limit of detection (LOD) of 200 virions/μL (Ct of ∼ 30) [9]. We hypothesized that routine RT-LAMP testing would significantly decrease the opportunity for workplace transmission of SARS-CoV-2. We endeavored on a 9 month-long prescreening program for laboratory staff and on occasion included household contacts.

**Figure 1.**
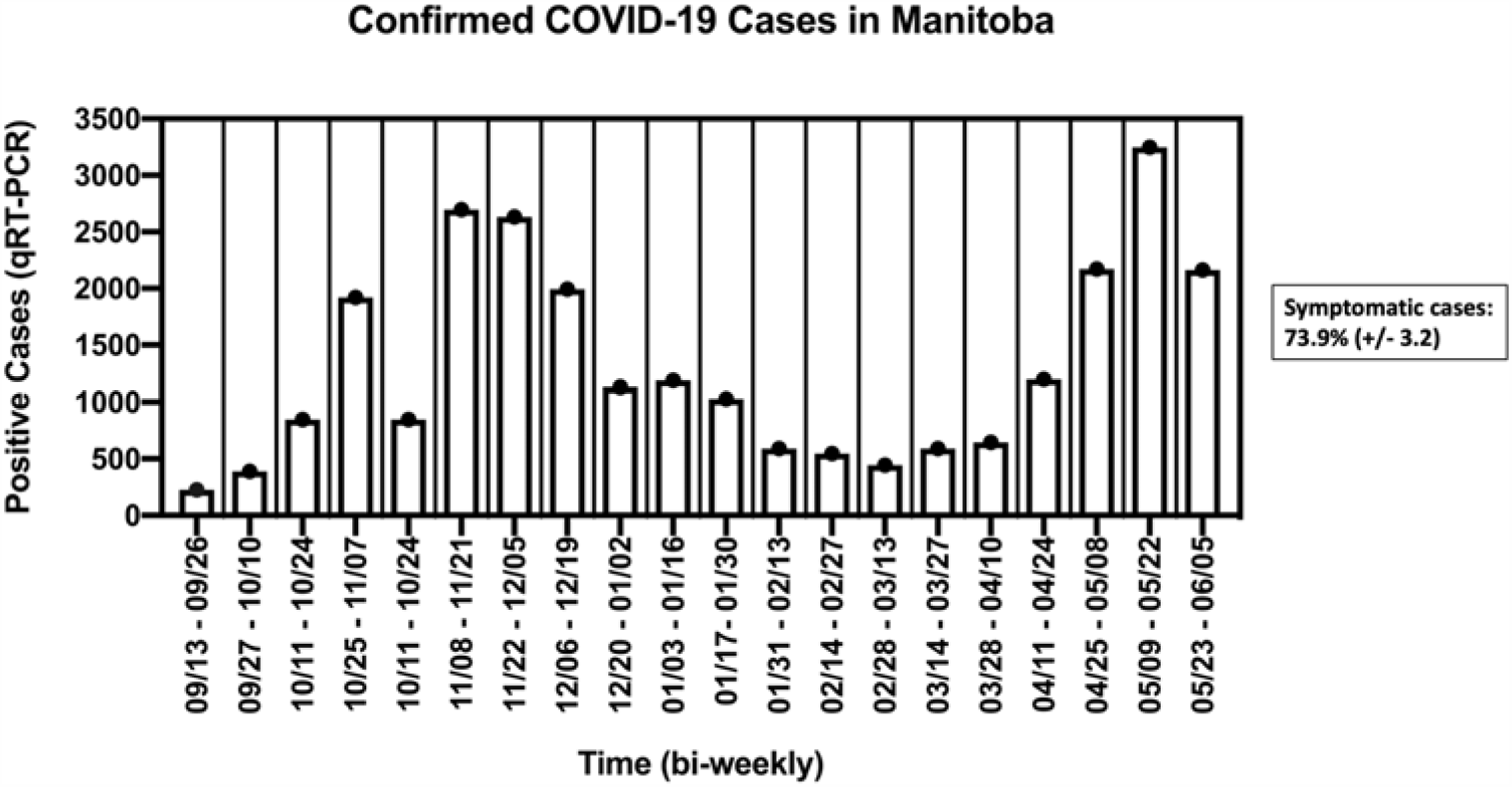
Epidemiological data of qRT-PCR positive cases in Manitoba from September 13, 2020 to June 5, 2021. Data was compiled from Manitoba’s Provincial COVID-19 Surveillance System (https://www.gov.mb.ca/health/publichealth/surveillance/covid-19/index.html) presented as a rolling average of two-week case counts. Every two-week interval included in the analysis reported the percentage of individuals which were symptomatic at the time of testing (73.9% (+/-3.2)). An overall average was calculated from each two-week interval.

## Materials and Methods

### Study Design

The premise of the testing regime was to prevent or reduce the likelihood of workplace transmission especially during times when high demand would result in qRT-PCR testing/results delays. Employees were asked to remain home if they or family members were sick and asked for voluntary collection of a contact-less saliva sample for testing. While at the workplace, non-pharmaceutical interventions such as physical distancing, masking and frequent handwashing were adhered to, in compliance with local public health orders [10]. A 3-testing-days-a-week (generally Monday, Wednesday, Friday and included Tuesday or Thursdays in lieu of an observed holiday) plan was implemented to voluntarily test employees prior to beginning work. Outlier samples from friends and family were sporadically accepted. Employees were given a unique identification number by random draw, this number was used throughout the testing period, then 2 to 4 employee samples were pooled as a group with a letter designation. Outlier samples were given a unique number and were either tested individually or were pooled with other outliers to comprise a unique and separate letter group. The program commenced on September 16, 2020 and concluded on May 31, 2021. Positive group pools were disaggregated and individually retested (referred to as reflex testing) with all three SARS-CoV-2 RT-LAMP primer sets to determine a true positive. Test-positive individuals were asked to leave the work-place and follow the Manitoba Public Health Guidelines regarding isolation and visit an official COVID-19 Testing Centre for qRT-PCR testing. Pending qRT-PCR confirmation of a positive result employees adhered to return to work policies set by Manitoba public health [11]. Figure 2 demonstrates expected results for the screening program.

**Figure 2.**
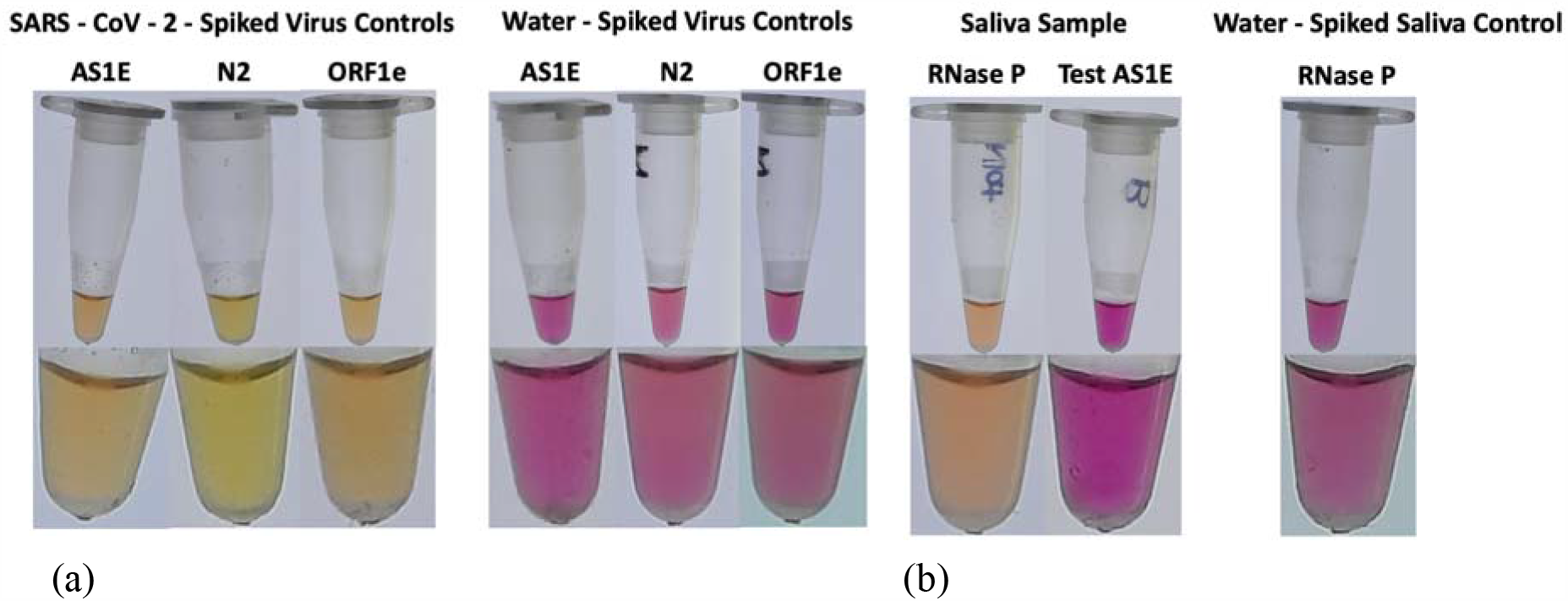
Expected RT-LAMP Reaction Results. (a) Control saliva-sample tubes were mixed 1:1 with Stabilization buffer and nuclease-free water, spiked with 2 μL of heat-inactivated SARS-CoV-2 or with 2 μL nuclease-free water, as indicated, and incubated for 10 minutes at 95°C. Four μL of respective mixtures were added to RT-LAMP reactions with SARS-CoV-2 primer sets (AS1E, N2 or ORF1e) and incubated for 30 minutes at 65°C followed by 2 minutes at 80°C. Images were taken on a lightbox with 4X (top panel) and 10X (bottom panel) magnification. (b) Saliva-sample tubes were mixed 1:1 with Stabilization buffer and saliva or nuclease-free water, as indicated, and incubated for 10 minutes at 95°C. RT-LAMP reactions with the RNase P primer set were prepared, four μL of the respective mixtures were added and incubated for 30 minutes at 65°C followed by 2 minutes at 80°C. Images were taken on a lightbox with 4X (top panel) and 10X (bottom panel) magnification. A negative amplification of the gene target results in the solution to remain pink (basic pH) whereas, target gene amplification results in a color change from pink to yellow (acidic pH).

### Saliva-sample preparation

Sample tube master mix preparation. Sample tubes were prepared with 2X Stabilization buffer, as described previously [9]. Briefly, a stock containing: 5mM (717 mg) TCEP-HCl (Gold Bio #TCEP10, Cedarlane Labs, Burlington, Canada), 2mM (2 mL) 0.5M pH8 EDTA (#324506-100ML, Millipore-Sigma, Oakville, Canada), 29mM (1.45 mL) 10M NaOH pellets (#S8045-500G, Millipore-Sigma, Oakville, Canada) previously dissolved in nuclease-free water, 100 μg/mL (50mg) recombinant PCR-grade Proteinase K (Roche #3115879001, Cedarlane Labs, Burlington, Canada) and then the solution was brought up to 500 mL with nuclease-free water (10220-404, VWR, Mississauga, Canada). One mL aliquots of 2X Stabilization buffer were deposited into nuclease-free, graduated, 5 mL screw-cap sample tubes (#10002-738, VWR, Mississauga, Ontario, Canada) and stored at 4°C until use. Saliva-sampling. On scheduled testing days, employees were asked to abstain from consuming any food, beverages or brushing teeth half an hour prior to producing a sample. The employees would independently remove a sample tube-containing 1 mL 2X Stabilization buffer from 4°C and provide 1 mL of fresh saliva from an isolated location: then tubes were shaken vigorously for 10 seconds, labeled with their respective unique number identifier, the exterior was decontaminated with 70% ethanol for 60 seconds at room temperature then stored at 4°C in a designated location [12], [13]. Samples were generally processed for the LAMP assay within a few hours; however, samples may be stored for up to 4 days [9].

### Saliva-sample processing

Sample tubes were vortexed briefly, then heat-inactivated in water baths (thermometer-verified) for 10 minutes at 95°C. Tubes were then plunged in ice and transferred into a biological safety cabinet for LAMP assay preparation.

### Colorimetric Reverse Transcription Loop-Mediated Isothermal Amplification (RT-LAMP) Assay

#### Primers

Desalted primer sets were fabricated by IDT (Coralville, Iowa, USA), according to the sequences provided previously by Yang et. al. [9]. Primer sets were diluted and mixed to make 10X concentration of primers targeting different regions in the SARS-CoV-2 genome: the ORF1ab gene (A1SE and ORF1e) or the N gene (N2) and, a control for human saliva (RNaseP) that detects the RNase P gene. A1SE, ORF1e, N2 and RNaseP primer sets contained: FIB and BIP (16 μM), LF and LB (4 μM) and F3 and B3 (2 μM) primers, except LF was omitted from N2. The sequences are provided in Supplementary Table S1.

#### RT-LAMP assay master mix

Reactions were prepared as independent master mixes as described previously [9]. Briefly, 4 μL of nuclease-free water, 10 μL Warmstart colorimetric LAMP 2X master mix (#M1800L, New England Biolabs, Whitby, Canada) and 2 μL respective 10X primer set mix (either AS1E, ORF1e, N2 or RNase P) were combined and aliquoted into PCR tubes. For routine testing only the AS1E reactions were used for randomly pooled saliva samples and virus controls and only 1 group at random was tested for RNAse P. For test reactions, either 4 μL from an individual’s saliva-stabilization buffer sample or an equal division of up to four saliva-stabilization samples was added but never exceeded four pooled samples (1 μL each) per group. Negative control and SARS-CoV-2 positive control reactions were prepared similarly, except four μL nuclease-free water in 2X stabilization buffer or 4 μL of SARS-CoV-2 control virus, were added instead of saliva. The 20 μL reactions were gently vortex and incubated in a thermal cycler at 65°C for 30 minutes, but never exceeded 35 minutes, followed by heat inactivation for 2 minutes at 80°C to preserve to color and prevent further unwanted amplification. Reaction tubes were visually inspected for color-change due to pH changes during amplification of viral RNA (basic pH-pink color: negative and acidic pH-yellow color: positive) and documented, immediately after heat inactivation. In the event of a positive test, the group was split into individual samples and tested with all primer sets along with the appropriate negative and virus controls.

#### SARS-CoV-2 virus controls

Two microliters from a stock of 1.6 × 10^5^ TCID_50_/mL (1.8 × 10^8^ RNA genome copies/mL) heat-inactivated SARS-CoV-2 (2019-nCoV/USA/WA1/2020, ATCC VR-1986HK, #2225411, Cedarlane Labs, Burlington, Canada) was aliquoted into nuclease-free 0.2 mL PCR tubes (Axygen Scientific PCR-02-C, #10011-780, VWR, Mississauga, Canada) equating to approximately 320 TCID_50_ units (3.6 × 10^5^ RNA genome copies) and stored at −80°C. An aliquot was added to a saliva-sample tube containing: 1 mL 2X stabilization buffer and 1 mL nuclease-free water (in place of saliva), heat-inactivated (10 minutes at 95°C), cooled on ice (5 minutes) and 4 μL were added to reaction tubes, approximately equivalent to 0.64 TCID_50_ units (720 RNA genome copies) of SARS-CoV-2.

## Results

During the 9-month screening program, n= 1,649 saliva samples were tested with 2 positive cases detected, both were from the same household (percent positivity: 0.12%). The epidemiological curve of reported COVID-19 positive cases in Manitoba during the study period is summarized in Figure 1. Two surges in observed cases occurred from November 8 to November 28 and May 9 to May 22 (Figure 1). During the November case surge, on the evening of a regular workday (herein referred to as “Day 1”, an employee, referred herein as “E” (who tested negative earlier that morning on the scheduled testing day) reported that a loved one had a “scratchy throat” (herein referred to as “F”). E was asked not to report for work on the following day (Day 2); however, E and F delivered samples contactless for non-scheduled testing. E tested negative, and F tested positive with the AS1E primer set. F was retested with appropriate controls and was found to be positive again with AS1E and with ORF1e, but not with N2 (Figure 3 (a)). Both E and F attempted to isolate from each other at home and immediately scheduled an appointment for qRT-PCR at a COVID-19 Testing Centre. However, since the earliest available was days later (Day 5), the decision was made that E should not report to work. On the next regularly scheduled testing day (Day 3), E again was negative, and F remained positive (Figure 3 (b)). On Day 4, a non-scheduled testing day, E became positive with the AS1E primer set and was not retested with any other primer sets. Additionally, F did not provide a sample (Figure 3 (c)). On Day 5, E and F were administered nasopharyngeal swabs and subsequently tested by qRT-PCR, both were confirmed positive 2 days later (Day 7). E and F complied with Manitoba Public Health’s recommendations and E returned to work when the isolation period had officially ended on Day 17, as summarized in Figure 4.

**Figure 3.**
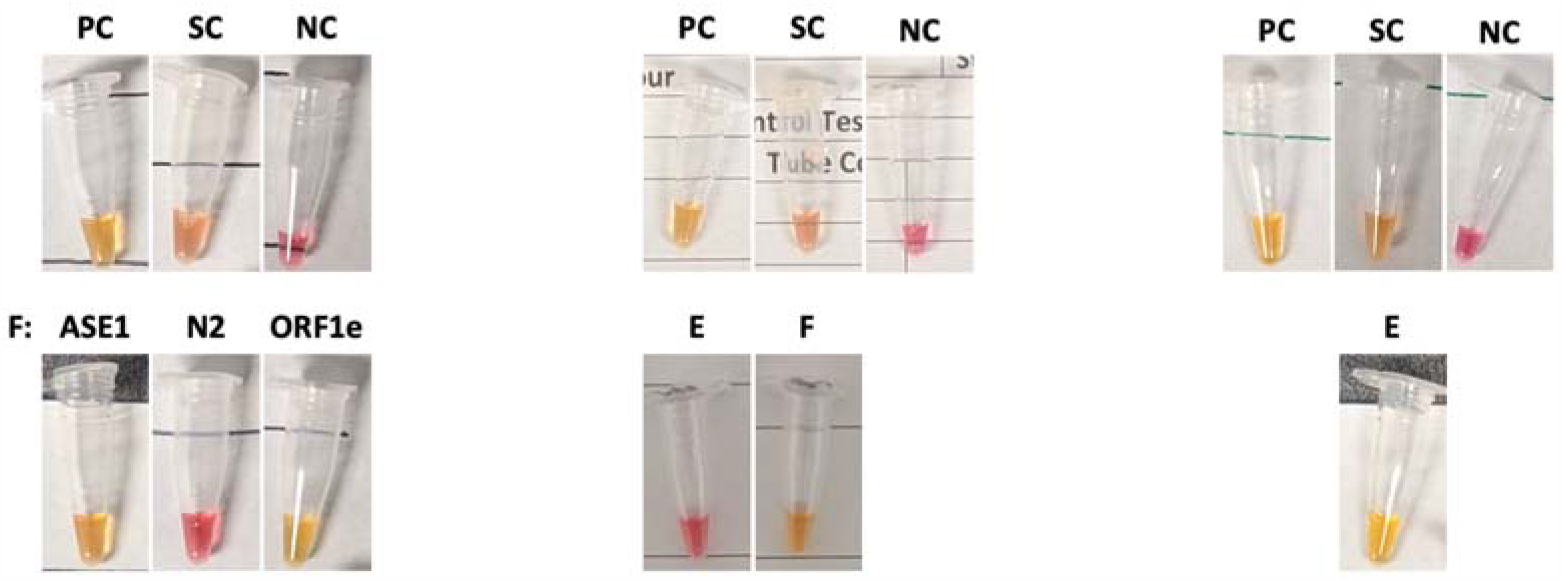
Original RT-LAMP Test Results from Individual’s E and F. (a) Day 2: individual F’s saliva sample processed and subjected to RT-LAMP using AS1E, N2 and ORF1e primer sets against SARS-CoV-2. (b) Day 3: individual E and F’s saliva were tested using AS1E per a regular scheduled testing day. (c) Day 4: individual E’s saliva sample was deemed positive with AS1E primer set against SARS-CoV-2. PC – SARS-CoV-2 virus control, SC – Saliva control (RNase P), NC – Negative control, AS1E, N2 and ORF1e – primer sets targeting SARS-CoV-2 genes. A negative amplification of the gene target results in the solution to remain pink (basic pH) whereas, target gene amplification results in a color change from pink to yellow (acidic pH).

**Figure 4.**
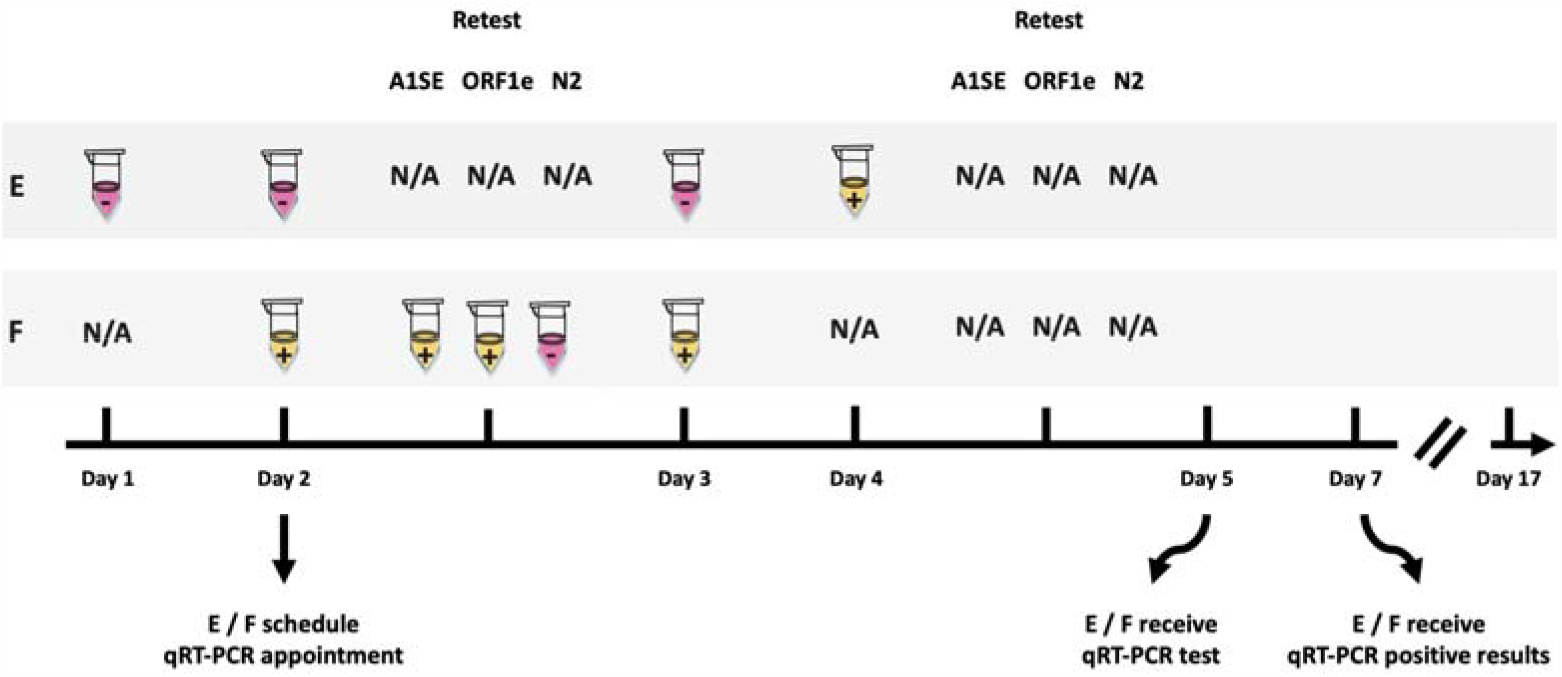
Timeline of E and F cases. Beginning on Day 1, individual E tested negative for SARS-CoV-2. Day 2 saliva samples for E and F were subjected to RT-LAMP using AS1E. Upon F testing positive, the sample retested positive with AS1E and ORF1e but not for the N2 primer sets against SARS-CoV-2. E and F scheduled an appointment for qRT-PCR testing. On Day 3, E tested negative, and F remained positive. On Day 4, E’s saliva sample was positive for SARS-CoV-2 with the AS1E primer set and further testing was omitted. Day 5 and Day 7: E and F received qRT-PCR tests and were confirmed positive, respectively. A positive test for SARS-CoV-2 is represented in yellow with a “+” and a negative test is denoted in pink with “-”.

## Discussion

We describe a simple and cost-effective program that can be applied in the workplace or other private setting to screen for SARS-CoV-19 infection and recommend employees or their close contacts for clinical diagnostic testing. The results from screening (around 15 to 20 samples per test day) employees and their friends and family members, demonstrate that frequent testing can identify infectious individuals and limit the spread of SARS-CoV-2. Although during the 9-month program, only 2 samples were positive for SARS-CoV-2, public health measures in Manitoba cycled between aggressive lockdowns and slightly relaxed restrictions. Undoubtedly, public health interventions played a role in the decreased case prevalence. In retrospect, given the increasing caseloads, one could have anticipated higher case numbers in the workplace, especially during the epidemiological week of the 2 positive samples (November 1, 2020 to November 7, 2020). The province’s testing and positivity rate increased by 1.1%, a change of n=1,451 administered tests and n=1,019 more positive cases were observed compared to 1 incubation period earlier (October 18 - 24, 2020) [14], [15]. Moreover, the delays in testing (3 days) and results (2 days) as experienced by E and F could have led to transmission in the workplace and possibly at employee residences or in public settings.

The choice to collect saliva specimens instead of nasal or nasopharyngeal swabs include ease-of-use (self-collection), enhanced safety of testing personnel and an increased likelihood of compliance. Numerous reports corroborate that molecular detection of SARS-CoV-2 from saliva samples does not significantly differ from the diagnostic accuracy of nasopharyngeal swabs [16]–[18] especially during the prodromal phase or in asymptomatic individuals [16], [19]. A scheme that prioritizes saliva over swabs would be more amenable for routine testing in children. This would be very powerful as children, despite tending to display milder symptoms or remaining asymptomatic when compared to adults [20], may shed infectious virus at similar levels [21], [22].

Direct saliva testing with the Warmstart product in relation to hydrolysis-probe qRT-PCR, demonstrated a limit of detection (LOD) of 200 virions/μL (Ct of ∼ 30) [9] and 100-1000 viral RNA copies (in vitro transcribed RNA) (Ct of ∼ 30.2 (+/-2.6)). Thus, Warmstart appears to be 10 to 100 times less sensitive than qRT-PCR [23]. Of note, the sensitivity of RT-LAMP can be improved using alternative enzymes, reagents, dye preparations and an optional RNA extraction step, as demonstrated after the conclusion of our study [23]. Differences in sensitivity between RT-LAMP and qRT-PCR has been shown in to be comparable when testing frequency was daily, every three and every seven days in epidemiologic modeling [5]. Thus, the strategy of increased testing frequency can overcome lower sensitivity with the additional benefits of: non-invasive sample collection, lower cost, availability, and consistency when detecting highly contagious individuals (Ct < ∼ 30 – 32) in real-world settings [9], [23]. It is worth noting that these authors used different primer sets for their RT-LAMP studies and that Ct values may vary based on the choice of hydrolysis-probes, reagent chemistries and machine variability.

Variants were not documented by the Manitoba government until the week of February 7 to February 13, 2021 (epidemiological week 6) from then onwards to the end of the study, the predominant variants were: either “undefined” or were B.1.1.7. (Alpha) in approximately 5 - 30% of cases depending on the week [24]. Using the Global Initiative on Sharing Avian Flu Data (GISAID) initiative, we compared the primer set sequence fidelity (AS1E, ORF1e and N2) to the first variant described during the study period. In chronological order the Beta, Alpha, Gamma and Delta variants were identified on samples collected on January 21, 2021 (accession: EPI_ISL_1594283), January 16, 2021 (accession: EPI_ISL_1594098), March 31, 2021 (accession: EPI_ISL_1594071) and April 29, 2021 (accession: EPI_ISL_2495627), respectively (Supplementary Table S2). Additionally, the Lambda and Mu variants were not detected prior to June 1, 2021. The AS1E primers remained identical to Alpha, Beta, Gamma and Delta. However, the F3 from the ORF1e set had a single nucleotide change (C to T) in Alpha only. More importantly, the N2 primer set had two primers: Loop B and FIP that had multiple changes. Loop B had a single difference (G to T) when compared to Delta. Whereas FIP demonstrated 3, 5 and 1 nucleotide changes for Alpha, Gamma and Delta, respectively (Supplementary Table S2). In the context F’s saliva sample testing negative with N2: even though Alpha was not documented until January, it is unclear if Alpha may have been circulating during late October. While it is tempting to speculate that an infection with Alpha may have caused this failure, it is more likely a false negative.

The potential for false negative reporting is a limitation of the study: i) screening for a at least 2 or 3 gene targets instead of 1 (AS1E) would decrease the likelihood of a false negative. ii) RNaseP detection was only performed in 1 group per scheduled testing day. It is possible that some saliva samples may be lacking an adequate concentration of buccal cells for each group, indicating poor sample collection.

## Conclusions

During a nine-month, voluntary SARS-CoV-2 testing program, two positive samples out of nearly n= 1700 were identified and confirmed by subsequent tests. This type of program helps disseminate the knowledge that inexpensive, convenient, primers can be adjusted for new variants for routine rapid testing for SARS-CoV-2. These assays can be accomplished in under an hour with minimal expertise and with reduced risks to the test administrator in multiple settings including, workplaces and schools.

## Data Availability

All data produced in the present study are available upon reasonable request to the authors

## Supplementary Materials

Table S1: primer sets for RT-LAMP assay and nucleotide changes observed in variants, Table S2: GISAID sequences used in study.

## Author Contributions

BWMC and SST conceived and designed the testing program. DGP coordinated ordering of supplies and generating protocols. BWMC and DGP optimized the RT-LAMP assay including virus controls. KK, MT, and NT performed the routine RT-LAMP testing and results documentation. BWMC interpreted data and wrote the manuscript. All authors provided feedback and editing. All authors have read and agreed to the published version of the manuscript.

## Author Disclaimer

Conclusions or learnings were not gained through research (for wide external application) but through a QI/QA project carried out in the local context.

## Institutional Review Board Statement

Ethical review and approval were waived for this study, due to a non-human subjects research determination and ethical evaluation which was deemed ineligible for Institutional Review Board review by Sterling IRB, protocol number 9576.

## Informed Consent Statement

Informed consent was obtained from all subjects involved in the study. Written informed consent has been obtained from the patient(s) to publish this paper.

## Data Availability Statement

Manitoba’s epidemiological data can be obtained from the “Epidemiological & Surveillance Unit” provided by the Government of Manitoba (https://www.gov.mb.ca/health/publichealth/surveillance/covid-19/index.html). Positive test case data from routine testing is contained within the manuscript (Figure 1, Figure 3 and Figure 4) and in supplementary material (Table S1 and Table S2). Data available on request due to privacy and ethical restrictions. Official testing results documents presented in this study are available on request from the corresponding author. The data are not publicly available due to the privacy of the documents.

## Acknowledgments

We would like to acknowledge the collection and submitting laboratories which shared SARS-CoV-2 variant sequences with GISAID (Supplementary Table S2).

## References

1. M. J. Mina, R. Parker, and D. B. Larremore, “Rethinking Covid-19 Test Sensitivity — A Strategy for Containment,” N. Engl. J. Med., p. NEJMp2025631, Sep. 2020, doi: 10.1056/NEJMp2025631.

2. C. MacLean, “With days-long waits for results, some Winnipeg parents question value of getting COVID-19 tests,” Canadian Broadcasting Corporation (CBC) News, 2020, Oct. 17, 2020.

3. J. Keele, “Results for COVID-19 test times seeing delays as cases grow in Manitoba,” Canadian Television Network (CTV) News, May 04, 2021.

4. P. Lam, “Frustrated Manitobans say they’re waiting days for COVID-19 test results,” Canadian Broadcasting Corporation (CBC) News, Winnipeg, Oct. 07, 2020.

5. D. B. Larremore et al., “Test sensitivity is secondary to frequency and turnaround time for COVID-19 screening,” Sci. Adv., vol. 7, no. 1, pp. 1–10, 2021, doi: 10.1126/sciadv.abd5393.

6. M. A. Johansson et al., “SARS-CoV-2 Transmission from People without COVID-19 Symptoms,” JAMA Netw. Open, vol. 4, no. 1, pp. 1–8, 2021, doi: 10.1001/jamanetworkopen.2020.35057.

7. A. L. Rasmussen and S. V. Popescu, “SARS-CoV-2 transmission without symptoms,” Science (80-.)., vol. 371, no. 6535, pp. 1206–1207, 2021, doi: 10.1126/science.abf9569.

8. T. Notomi et al., “Loop-mediated isothermal amplification of DNA.,” Nucleic Acids Res., vol. 28, no. 12, p. E63, Jun. 2000, doi: 10.1093/nar/28.12.e63.

9. Q. Yang et al., “Saliva TwoStep for rapid detection of asymptomatic SARS-CoV-2 carriers,” Elife, vol. 10, no. e65113, pp. 1–28, 2021, doi: 10.7554/eLife.65113.

10. M. Government, “General COVID-19 Prevention Orders,” Winnipeg, 2020. doi: 10.1016/S0140-6736(02)63253-2.

11. P. H. Manitoba, “COVID-19 Novel Coronavirus Guidelines: Key Responsiblities of Employees, Managers and Employers,” 2020. [Online]. Available: https://www.gov.mb.ca/asset_library/en/coronavirus/workplace_responsibilities.pdf.

12. G. Xiling et al., “In vitro inactivation of SARS-CoV-2 by commonly used disinfection products and methods,” Sci. Rep., vol. 11, no. 1, pp. 1–9, 2021, doi: 10.1038/s41598-021-82148-w.

13. A. Kratzel et al., “Inactivation of Severe Acute Respiratory Syndrome Coronavirus 2 by WHO-Recommended Hand Rub Formulations and Alcohols,” Emerg. Infect. Dis., vol. 26, no. 7, pp. 1592–1595, 2020, doi: 10.3201/eid2607.200915.

14. M. Government, “Provincial COVID-19 Surveillance,” Winnipeg, 2020. Accessed: Nov. 30, 2021. [Online]. Available: https://www.gov.mb.ca/health/publichealth/surveillance/covid-19/2020/week_45/index.html#epi_curve.

15. M. Government, “Provincial COVID-19 Surveillance,” Winnipeg, 2020. Accessed: Nov. 30, 2021. [Online]. Available: https://www.gov.mb.ca/health/publichealth/surveillance/covid-19/2020/week_43/index.html.

16. S. Iwasaki et al., “Comparison of SARS-CoV-2 detection in nasopharyngeal swab and saliva.,” J. Infect., vol. 81, no. 2, pp. e145–e147, 2020, doi: 10.1016/j.jinf.2020.05.071.

17. S. Wei et al., “Field-deployable, rapid diagnostic testing of saliva samples for SARS-CoV-2,” medRxiv Prepr. Serv. Heal. Sci., 2020, doi: 10.1101/2020.06.13.20129841.

18. K. Zhang, A. Shoukat, W. Crystal, J. M. Langley, A. P. Galvani, and S. M. Moghadas, “Routine saliva testing for the identification of silent coronavirus disease 2019 (COVID-19) in healthcare workers,” Infect. Control Hosp. Epidemiol., vol. 42, no. 10, pp. 1189–1193, 2021, doi: 10.1017/ice.2020.1413.

19. R. Wölfel et al., “Virological assessment of hospitalized patients with COVID-2019,” Nature, vol. 581, no. 7809, pp. 465–469, 2020, doi: 10.1038/s41586-020-2196-x.

20. M. Hippich et al., “Public health antibody screening indicates a six-fold higher SARS-CoV-2 exposure rate than reported cases in children,” Med, 2020, doi: 10.1016/j.medj.2020.10.003.

21. L. M. Yonker et al., “Virologic Features of Severe Acute Respiratory Syndrome Coronavirus 2 Infection in Children.,” J. Infect. Dis., vol. 224, no. 11, pp. 1821–1829, Dec. 2021, doi: 10.1093/infdis/jiab509.

22. T. Jones et al., “An analysis of SARS-CoV-2 viral load by patient age,” 2020, doi: 10.1101/2020.06.08.20125484.

23. C. Amaral et al., “A molecular test based on RT-LAMP for rapid, sensitive and inexpensive colorimetric detection of SARS-CoV-2 in clinical samples,” Sci. Rep., vol. 11, no. 1, pp. 1–12, 2021, doi: 10.1038/s41598-021-95799-6.

24. M. Government, “Provincial COVID-19 Surveillance,” Winnipeg, 2021. Accessed: Nov. 28, 2021. [Online]. Available: https://www.gov.mb.ca/health/publichealth/surveillance/covid-19/index.html.

