## Supplemental Table 1 for "Implementation of a Rapid RT-LAMP Saliva-based SARS-CoV-2 Testing Program in the Workplace"

**Supplementary Table:**

**Table S1: Primer Sets For RT-LAMP Assay and Nucleotide Changes Observed in Variants.**

| Primer Set | Primer Name | Primer Sequence (5’-3’) | Alpha | Beta | Gamma | Delta |
| --- | --- | --- | --- | --- | --- | --- |
| AS1E | F3 | CGGTGGACAAATTGTCAC | NC | NC | NC | NC |
|  | B3 | CTTCTCTGGATTTAACACACTT | NC | NC | NC | NC |
|  | Loop F | TTACAAGCTTAAAGAATGTCTGAACACT | NC | NC | NC | NC |
|  | Loop B | TTGAATTTAGGTGAAACATTTGTCACG | NC | NC | NC | NC |
|  | FIP | TCAGCACACAAAGCCAAAAATTTATTTTTCTGTGCAAAGGAAATTAAGGAG | NC | NC | NC | NC |
|  | BIP | TATTGGTGGAGCTAAACTTAAAGCCTTTTCTGTACAATCCCTTTGAGTG | NC | NC | NC | NC |
| N2 | F3 | CGGCAGTCAAGCCTCTTC | NC | NC | NC | NC |
|  | B3 | TTGCTCTCAAGCTGGTTCAA | NC | NC | NC | NC |
|  | Loop F | This primer was omitted from the set. (Yang et al. 2020) | NA | NA | NA | NA |
|  | Loop B | ATGGCGGTGATGCTGCTCTT | NC | NC | NC | 1 (G-->T^*^) ATGGCTGTGATGCTGCTCTT |
|  | FIP | TCCCCTACTGCTGCCTGGAGCGTTCCTCATCACGTAGTCG | 3 (CCC-->GTT^*^) TCGTTTACTGCTGCCTGGAGCGTTCCTCATCACGTAGTCG | NC | 5 (CCC -->GTT^*^, CT-->GA^*^) TCGTTTAGAGCTGCCTGGAGCGTTCCTCATCACGTAGTCG | 1 (C-->A^*^) TCCCATACTGCTGCCTGGAGCGTTCCTCATCACGTAGTCG |
|  | BIP | TCTCCTGCTAGAATGGCTGGCATCTGTCAAGCAGCAGCAAAG | NC | NC | NC | NC |
| ORF1e | F3 | GGCTAACTAACATCTTTGGC | 1 (C-->T^*^) GGCTAACTAATATCTTTGGC | NC | NC | NC |
|  | B3 | GTCAGCACACAAAGCCAA | NC | NC | NC | NC |
|  | Loop F | TCTTCAAGCCAATCAAGGAC | NC | NC | NC | NC |
|  | Loop B | TTGTCGGTGGACAAATTGT | NC | NC | NC | NC |
|  | FIP | TCTCTAAGAAACTCTACACCTTCCTTTTTACTGTTTATGAAAAACTCAAACC | NC | NC | NC | NC |
|  | BIP | TATCTCAACCTGTGCTTGTGAAATTTTAGAATGTCTGAACACTCTCCT | NC | NC | NC | NC |
| RNase P | F3 | TTGATGAGCTGGAGCCA |  | | | |
|  | B3 | CACCCTCAATGCAGAGTC |  |  |  |  |
|  | Loop F | ATGTGGATGGCTGAGTTGTT |  |  |  |  |
|  | Loop B | CATGCTGAGTACTGGACCTC |  |  |  |  |
|  | FIP | GTGTGACCCTGAAGACTCGGTTTTAGCCACTGACTCGGATC |  |  |  |  |

^*^ Change observed in variant
