## Supplemental Table 2 for "Implementation of a Rapid RT-LAMP Saliva-based SARS-CoV-2 Testing Program in the Workplace"

**Supplementary Table:**

**Table S2: GISAID Sequences Used in Study.**

| Variant | Isolate | Accession ID | Collection Date | Submission Date |
| --- | --- | --- | --- | --- |
| Alpha | hCoV-19/Canada/MB-NML-16346/2021 | EPI_ISL_1594283 | 2021-01-21 | 2021-04-15 |
| Beta | hCoV-19/Canada/MB-NML-17441/2021 | EPI_ISL_1594098 | 2021-01-16 | 2021-04-15 |
| Gamma | hCoV-19/Canada/MB-NML-34961/2021 | EPI_ISL_1594071 | 2021-03-31 | 2021-04-15 |
| Delta | hCoV-19/Canada/MB-NML-70550/2021 | EPI_ISL_2495627 | 2021-04-29 | 2021-06-11 |

All samples were collected from MB-Cadham Laboratory and submitted by the National Microbiology Laboratory (NML) in North America, Canada, Manitoba by: Anna Majer; Anneliese Landgraff; CanCOGeN’s metadata curation team; Darian Hole; David Alexander; Elsie Grudeski; Gary Van Domselaar; Grace Seo; Jared Bullard; Jennifer Tanner; Kerry Dust; Kirsten Biggar; Madison Chapel; Morag Graham; Natalie Knox; Nathalie Bastien; Paul Van Caeseele; Philip Mabon; Public Health Agency of Canada CanCOGeN team; Rhiannon Huzarewich; Russell Mandes; Shari Tyson; Timothy Booth; Yan Li.
